# Neurophysiological response and connectivity changes in the STN with a month-long exercise intervention

**DOI:** 10.1101/2025.07.15.25331621

**Authors:** Prajakta Joshi, Lara Shigo, Brittany Smith, Camilla W. Kilbane, Aratrik Guha, Amit K. Sinha, Kenneth A. Loparo, Angela L. Ridgel, Aasef G. Shaikh

## Abstract

Parkinson’s disease (PD) affects over 1 million people in the U.S., posing challenges in symptom management. Treatments include medications like levodopa, surgical options such as deep brain stimulation (DBS), and rehabilitative approaches like exercise. While the effects of levodopa and DBS on basal ganglia circuits—especially the subthalamic nucleus (STN)—are well studied, exercise-induced changes in STN activity remain underexplored. This study investigates the short-and long-term effects of a month-long motorized cycling intervention on STN activity, using local field potentials (LFPs) recorded from 29 electrodes across 18 STNs in 9 participants.

Consistent with our previous work, dorsal STN regions showed stronger 1/f noise, peak oscillatory power, and 8–35 Hz activity than ventral regions. Acute effects were minimal, but long-term data revealed increased peak and alpha/beta power in dorsal STN—distinct from the beta suppression seen with levodopa or DBS, suggesting a unique exercise-driven modulation.

We further analyzed dorsal–ventral interactions using phase slope index (PSI) and imaginary coherence (iCOH). Low PSI (< 0.1) indicated minimal crosstalk, implying independent dorsal and ventral oscillators. However, iCOH showed beta coherence initially rose significantly over time. Using a novel System Structure (SStr) algorithm, we assessed causality between dorsal and ventral signals. Preliminary findings suggest both are influenced by a hidden driver, likely the exercise. In summary, exercise induces sustained, distinct changes in STN activity, possibly mediated by extrinsic inputs acting in parallel on dorsal and ventral regions. Further studies are needed to clarify these mechanisms.

## Introduction

Parkinson’s Disease (PD) affects over 1 million people in the United States and 8 million worldwide (Dorsey et al., 2018; Marras et al., 2018). It results from the loss of dopaminergic neurons in the substantia nigra, a critical region within the basal ganglia (BG) circuitry responsible for motor control (Colcher and Simuni, 1999). Pharmacotherapy, particularly with levodopa (Cotzias et al., 1969), is effective in the early stages as it offers rapid relief from symptoms like tremor and rigidity but has variable effects on gait and balance (Pirker et al., 2023; Smulders et al., 2016). Deep brain stimulation (DBS), typically employed in advanced stages, reduces medication dependence and effectively manages motor symptoms such as tremor and rigidity (Groiss et al., 2009). In addition to medication and surgical interventions, exercise plays a pivotal role in PD rehabilitation. Its long-term benefits on gait, balance, and motor symptoms are well-established, as it promotes gradual physiological changes that enhance overall functionality and quality of life (Atterbury and Welman, 2017; Mak and Wong-Yu, 2019; Morris et al., 2010; Sparrow et al., 2016). Each treatment approach has distinct strengths and limitations, emphasizing the need to understand their underlying mechanisms. This study seeks to investigate the acute and long-term effect of exercise in the form of motor assisted stationary dynamic cycling on the neurophysiology of the basal ganglia.

Over the past two decades, numerous studies have investigated the short-term and long-term effects of exercise on various cellular-level markers of PD, particularly within BG and associated brain circuits. They have documented increases in structural proteins (like MAP2 and neurofilament), synaptic proteins (such as synaptophysin and synapsin I), neural activity markers (like Egr-1), and neurotrophic factors (such as glia-derived neurotrophic factor and brain-derived neurotrophic factor) (Garcia et al., 2012; Salame et al., 2016). In addition, neurotransmitters such as dopamine D2 receptor are increased in the striatum and glutamate receptors are decreased with exercise (Vučcković et al., 2010). Acute changes may differ significantly from those observed in long-term interventions (Salame et al., 2016). While the therapeutic effects of exercise are well-documented, most studies focus on molecular mechanisms, leaving the neurobiological basis largely unclear.

Imaging studies provide us with a macro level picture of dynamic changes in the brain due to exercise. Exercise in PD primarily leads to increased activation across multiple brain regions, including the cerebellum, occipital lobe, parietal lobe, and frontal lobe (Li et al., 2022). Forced biking exercise and anti-parkinsonian medication yield comparable improvements in PD symptoms and induce similar fMRI activation patterns (Alberts et al., 2016). A treadmill intervention combined with fMRI revealed alterations in interregional connectivity among structures such as the motor cortex, brainstem, prefrontal cortex, and subcortical regions, resulting in brain activity patterns that resembled those of healthy individuals (Ding et al., 2022). Rather than inducing isolated changes in a single brain area, exercise appears to promote functional enhancements through the coordinated involvement of multiple neural regions, potentially contributing to the observed improvements in motor and cognitive function (Silva et al., 2018). While Imaging studies offer a broad view of the brain’s response to exercise, neurophysiological studies provide high-temporal-resolution measurements of localized neural activity, including oscillatory patterns. Electroencephalogram studies enable recording from cortical layers, while structures placed deep inside the brain like the STN can be accessed for recording via DBS electrodes, which has been made possible with the new technological advances.

The neurophysiology of the STN has been extensively studied for its response to PD interventions like levodopa therapy and DBS. Dopamine depletion increases beta band (13-35 Hz) activity of the STN (Sharott et al., 2005). This beta hypersynchrony is reduced with dopamine replacement therapy drugs containing levodopa and dopamine agonists (Mathiopoulou et al., 2024; Priori et al., 2004). The changes in the beta band power also correlates with changes in motor symptoms (Kühn et al., 2006). DBS is also shown to suppress the beta power in the STN (Bronte-Stewart et al., 2009; Trager et al., 2016). Levodopa primarily suppresses the low-beta (13-20 Hz) activity and DBS reduces the entire beta band (Mathiopoulou et al., 2024). In this study, we have measured immediate and long-term STN neurophysiological changes in response to motor assisted stationary cycling with adaptive exercise planning. This type of high-cadence cycling has previously been shown to be linked to alleviating motor symptoms of PD (Kim et al., 2024; Ridgel et al., 2015; Ridgel and Ault, 2019). We also investigated connectivity and causal relationships within STN to understand the changes in the flow of information in response to biking.

## Methods

This study was performed in accordance with the Declaration of Helsinki. This human study was approved by the Kent State University and Cleveland VA Medical center institutional review boards. All adult participants provided written informed consent to participate in this study. The main objective of the study was to investigate potential physiological mechanisms of exercise effects in PD symptoms by examining its impact on STN neuronal activity. We analyzed STN local field potential (LFP) activity in nine PD patients (eight men and one woman; average age 66.4±9.7 years). The average Unified Parkinson’s Disease Rating Scale (UPDRS-III) score was 20.7±2.56, and the average daily dopamine dose was 1061.7±480 mg/day. Participants were included if they had a PD diagnosis confirmed by UK Brain Bank Criteria, a stable antiparkinsonian medication regimen for at least six months, bilateral STN deep brain stimulation (DBS) with Medtronic Percept™ PC implantable pulse generator and electrode lead models 3389 or Sense Sight B33005 Directional™ (Medtronic, Minneapolis, MN, USA), stable stimulation settings for at least one month, the ability to provide written informed consent, and the ability to perform exercise on a motorized stationary bike every other day for up to 12 sessions over four weeks. Those with cardiovascular risk factors or musculoskeletal injuries that would prevent safe participation in the exercise program were excluded.

### Exercise intervention

The intervention involved the use of a stationary motorized bicycle programmed to maintain a preset cadence of 80 revolutions per minute (RPM). The motor was responsible for driving the pedals at the specified speed and providing resistance to the participants, while they were instructed to exert a minimal amount of force to slightly exceed the motor-driven cadence. Although participants had the capacity to apply additional effort to pedal at a faster rate, the motor’s torque feedback was designed to resist changes in speed to sustain the designated cadence, thereby facilitating a dynamic interaction between the rider and the exercise bike. To ensure continuous engagement and prevent unilateral effort, the resistance was adjusted - either increased or maintained when participants were able to exceed the desired cadence (motor-driven speed). This interactive component was designed to optimize the neuromuscular engagement of the rider with the exercise bike. Real-time feedback on the interaction between the rider and the exercise bike was provided via a visual display. A comprehensive description of the mechanics underlying this dynamic cycling paradigm has been detailed in previous studies (Ridgel et al., 2017). A detailed account of the experimental paradigm is outlined in previous investigation from our lab (Kim et al., 2024).

### Experimental Setup

Participants underwent a structured motor assisted cycling-based exercise intervention that included up to 12 supervised sessions over four weeks, with each session spaced 48-72 hours apart. Cycling was performed on a custom-made motorized stationary bike at a cadence of 80 rotations per minute (RPM) for 30 minutes. Each session included a 5-minute warm-up and a 5-minute cool-down at 60 RPM (Figure 1A). The time interval between exercise and medication was kept consistent, and all sessions were performed at the same time of day. Participants were advised to avoid additional physical activities beyond their regular routine before the intervention to ensure that any observed changes were specifically due to the exercise intervention. Exercise intensity was monitored at two-minute intervals using a heart-rate monitor (Mi Band 6, Xiaomi, China or Polar H10 sensor, Polar, USA). Motor symptoms were measured using the KinesiaOne inertial measurement (IMU)-based device by Great Lakes Neurotechnology (Giuffrida et al., 2009; Mera et al., 2012) that scores the acquired IMU data before and after each exercise session on a scale of 0-4, 0 being normal movement and 4 designating maximum impact of PD on the movement. Motor tasks like involuntary movements (rest, postural and kinetic tremors), voluntary movements (upper and lower body movements) were included in the UPDRS examination. These symptoms were measured while ON-DBS. The rating of perceived exertion (RPE) was recorded every four minutes using the 6-20 Borg RPE scale. Out of the nine participants, six completed all 12 sessions. One participant dropped out of the study after the 9th session, and another after the 8th session, both due to unrelated seasonal flu-like illnesses. One participant dropped out of the study after the 11th session due to unrelated circumstances. Despite these disruptions, we were able to test our hypothesis with all participants included in the analysis.

**Figure 1:**
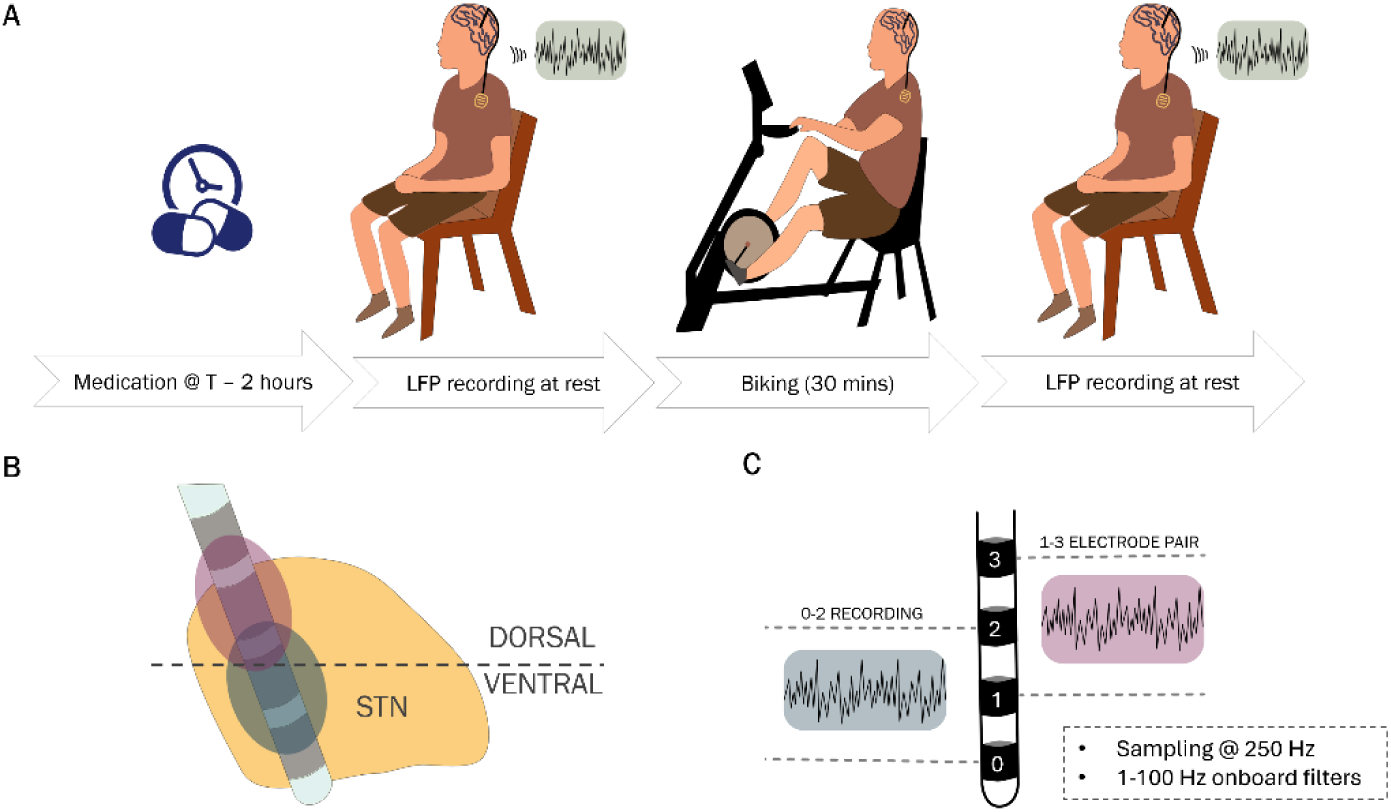
(A) Illustrates the steps followed during each of the 12 exercise sessions to obtain LFP recordings. The pre-biking LFP recording is conducted 120 minutes after the last medication, ensuring consistency in timing across sessions. The post-biking recording takes place after the participant has cycled for 30 minutes at 80 RPM, (B) Shows the positioning of the DBS lead within the STN. The dark gray dashed line represents the approximate division of the STN, indicating whether the electrode is positioned dorsally or ventrally, (3) Depicts the differential recordings obtained from Medtronic Percept PC system. Electrode pair 1-3 is classified as dorsal if electrode 2, positioned at the center, is located in the dorsal region, and as ventral if electrode 2 is in the ventral region. The same classification applies to electrode pair 0-2.

### LFP data acquisition

LFP data was recorded at 250Hz using the Medtronic Percept system in the indefinite streaming mode, with deep brain stimulation turned OFF. Recordings were conducted immediately before and after the exercise session while the participants were seated comfortably at rest with eyes open. Participants were instructed to remain silent and avoid any voluntary movements during the recording period. The same data collection procedure was followed for each session for every participant. DBS was turned off 10 minutes before every LFP data collection to eliminate any residual effects of stimulation. The recording was performed for a duration of 3.5 minutes (210 seconds). The on-board acquisition filter settings were set to 1Hz high-pass and 100Hz low-pass. The data was simultaneously recorded differentially from 3 electrode pairs from each brain hemisphere. The placement of electrodes varies among patients. To account for this variability and accurately interpret the results, it was necessary to identify the exact location of each recording pair within the STN. We used the pre-operative MRI and post-operative CT scans of our participants along with Lead-DBS toolbox (Horn and Kühn, 2015) to reconstruct the locations of the four contact pairs of interest for each patient. We then visually classified them into two categories: dorsal and ventral. As shown in Figure 1B, the electrodes placed in dorsal half of the STN were marked as dorsal, and the ones below the grey line were marked as ventral. The location of the grey line was marked by the axial plane passing though the center of the top dorsal and ventral boundary in the MNI coordinate space. Their locations were estimated based on the position of the center of the contact situated between the two differentially recording electrodes. For instance, the recording obtained from contact pair 0-2 was assigned using the location of contact 1, and the location of contact 2 was assigned to the recording obtained from contact pair 1-3, as depicted in Figure 1C. This process was repeated for all the contact pairs of interest in all our participants. Out of the 36 leads of interest (9 participants × 2 sides of the brain × 2 contact pairs of interest), we found 21 placed in the dorsal region of the STN, and 8 in the ventral region. Contacts found outside of the STN were not included in the analysis. Among the 29 electrodes, we identified 8 pairs placed within the same STN. These electrode pairs enabled coherency analysis between the two sites to examine changes in communication within the STN.

### LFP data analysis

The data was analyzed using custom MATLAB scripts (Version 2023b, MathWorks, USA) and the FieldTrip toolbox (Oostenveld et al., 2011). Following visual inspection of the electrophysiological data, instances of movement artifacts were identified and excluded from the dataset. Cardiac artifacts observed in two participants were removed using the template matching method implemented in the open source Perceive toolbox (https://www.github.com/neuromodulation/perceive/) (Merk et al., 2022). The processed data was then saved for further analysis. We performed spectral analysis to calculate power in different frequency bands such as alpha (8-12 Hz), low-beta (13-20 Hz), high-beta (21-35 Hz). To account for 1/f noise present in the neural spectrum recordings, we implemented the FOOOF algorithm to extract features such as the aperiodic exponent and peak power (Donoghue et al., 2020). To examine changes in the relationship between dorsal and ventral STN locations in response to exercise, we computed the imaginary part of coherence (iCOH) and the phase slope index (PSI) to assess communication within the STN (Ewald et al., 2012; Nolte et al., 2008). To examine the causal relation between the dorsal and ventral LFP data, we used the SStr algorithm, specifically Estimate 1 for identifying the existence of a hidden driver as provided in (Sinha and Loparo, 2021).

### Absolute and relative spectral power calculations

Power spectral density (PSD) curves were obtained for each contact by dividing the data in 1 second windows (1Hz frequency resolution) with 50% overlap. To account for within-subject variability and correct for impedance at different times, we normalized each PSD with the power of the 61-90Hz frequency band. Then, the absolute power was calculated by integrating the canonical frequency bands of alpha (8-12Hz), low-beta (13–20Hz), high-beta (21-35Hz), low-gamma (36-60 Hz) of the resulting curve. Relative power was calculated by dividing each of these absolute values by the cumulative sum of all PSD values in the 4 to 100 Hz region.

### Aperiodic spectrum and peak power extraction using FOOOF

The absolute PSD data was then processed with the FOOOF algorithm with the following parameters: frequency range of 8-50Hz, peak width limits of 2-20Hz, maximum number of peaks = 6, minimum peak height of 0, peak threshold of 2, and aperiodic model set to fixed. After computing the aperiodic fit using these parameters, we subtracted the aperiodic values from the full PSD to obtain the periodic PSD and recorded the aperiodic exponent values. The maximum value of the remaining spectrum was marked as the peak power of the spectrum, and the corresponding frequency was marked as the peak frequency.

### Imaginary part of coherence computation

Imaginary part of the coherence (iCOH) is a measure to quantify functional connectivity between two signals, specifically focusing on or isolating the interactions that are not influenced by shared noise or volume conduction. To quantify the iCOH values, we first obtained the coherency values for each signal pair using Equation *(1)* where C*_ij_* is the coherency between signals *i* and *j* at frequency *f*. S*_ij_* is the cross-spectral density between signals *i* and *j* at frequency *f.* S*_ii_* is the auto-spectral density of signal *i* at frequency *f*. S*_jj_* is the auto-spectral density of signal *j* at frequency *f*. iCOH at a given frequency is the imaginary component of the coherency value, as defined in Equation *(2)*. The significance of the iCOH values was assessed using the jackknife method, with a threshold of 2 indicating significance. We compute the iCOH values for interactions between 8 dorsal-ventral contact pairs and compare these values across different frequency bands.

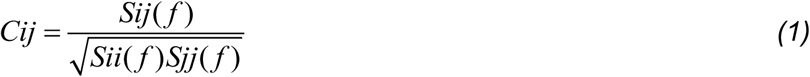

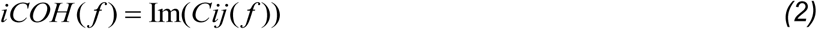

### Phase slope index (PSI) computation

PSI is a measure to quantify the directionality of interactions between two signals based on their phase relationships across frequencies. The value of PSI can range from-1 to 1. The Phase Slope Index (PSI) effectively captures relative time delays between different signals, aligning closely with the classical definition of linear phase spectra as defined in Equation *(3)*. It is robust against interference from non-interacting signals, regardless of their spectral content or superposition, ensuring that only meaningful interactions are considered. Additionally, PSI appropriately weights different frequency regions based on their statistical significance, making it a reliable tool for analyzing the temporal dynamics of signal interactions, where *ψ _ij_* = PSI, denotes the imaginary part and *F* denotes the set of frequencies under consideration in equation *(3)*. The PSI values are calculated between the dorsal-ventral pairs to understand the directionality of information exchange. The significance of the PSI values was assessed using the jackknife method, with a threshold of 2 indicating significance.

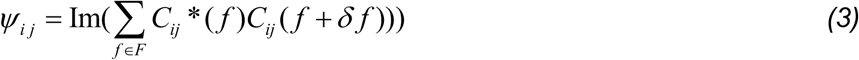

### Statistical analysis

After checking for normality of the distribution using histograms and q-q plots, the Wilcoxon signed-rank comparison was implmented to compare power in the frequency bands. For acute comparisons, the non-parametric Wilcoxon signed-rank test (WSRT) was used to compare power in frequency bands, iCOH and PSI values in a given range. A p-value of 0.05 was the criteria for significance in all statistical calculations. A Bonferroni correction was applied to the data derived from a single location and originating from the same calculation.

### Model for Long-Term Power Analysis

To establish if the LFP features and motor symptoms change systematically over time, we used a Generalized Linear Mixed Model (GLMM) with response variable – “Response” and the fixed effect of progressing sessions – “Session”, while accounting for variability across electrodes. The model included random intercepts and slopes for “Session” within each electrode, allowing both the baseline response and the effect of the session to vary across electrodes. The model was specified as - *Response∼Session+(1+Session∣Electrode)*

A gamma distribution with a log link function was employed, which is suitable for positively skewed data, to model the variable. The model was fitted using MATLAB’s fitglme function that uses maximum likelihood estimation for parameter estimation. Model diagnostics, including residual analysis and evaluation of random-effects variance components, were conducted to ensure the appropriateness of the model fit.

System Structuring for Causality Estimation: To estimate the causal relationship betweem the dorsal and ventral electrode data, the System Structuring (SStr) method (see Sinha and Loparo, 2021 and the references therein) was used. SStr is a set-theoretic, distribution-free, computational model for complex systems that quantifies, through a non-parametric network-diffusion procedure, the significance of components and inter-component interactions (both linear and non-linear). SStr computes a network model of a complex system *S* given by *S = (N, A, W_A_, W_N_)* where *N* is a set of system objects represented by nodes, *A* is a set of arcs interconnecting the system objects/nodes, and *W_A_* and *W_N_* are arc and node system measures, respectively. Subsystem *S_i_* of system *S* is given by the collection of nodes and arcs of *S* that correspond specifically to *S_i_*. The SStr network model of complex systems is able to identify and evaluate distinct distinguishing characteristics of the system, including causal relationships between subsystems, without requiring restrictive a-priori assumptions.

In this application, we are using SStr to estimate the causal relationship between the dorsal and ventral subsystems using the LFP time-series data recorded from the dorsal and ventral regions before and after the exercise intervention. A SStr model was developed using the data and the inter-system measures between composite dorsal and ventral subsystems and the cohesions of the individual subsystems were used to estimate the causal relationship between the subsystems. Cohesion is the mean self-connectivity over the system components, and the temporal variability of these system characteristics over time are indicative of variation in system interconnectivity and the direction of causality, Table 2.

## Results

This study aimed to examine neurophysiological changes in PD patients in response to exercise therapy, focusing on spectral characteristics and connectivity and causality within the STN*, and to identify acute, long-term, or adaptive changes occurring during the intervention*. We performed a comprehensive analysis of motor symptoms, spectral features such as power in canonical frequency bands (alpha, low beta, high beta), relative power, aperiodic exponent and peak oscillatory power in the LFP spectrum and strength and direction of causal interactions between the dorsal and ventral regions using data collected from 9 PD patients. The imaginary part of coherence, the phase slope index, and SStr interconnectivity measure and cohesion were used to analyze signaling between dorsal and ventral electrodes within the STN.

### Immediate and Long-Term changes in motor symptoms

KinesiaOne scores were used to quantify tremor and bradykinesia symptoms in the upper extremity. We combined resting, postural, and kinetic tremors on both sides – right and left to derive a combined tremor score. A reduction in tremor severity was observed in both acute (Figure 2A) and long-term recordings (Figure 2B) during biking. The effect was significant when comparing tremor scores immediately before and after exercise, with a slight but significant reduction from a median of 0.28 (IQR: 0.18–0.41) to 0.26 (IQR: 0.15– 0.36) following biking (WSRT, p = 0.007). Over a more extended period, total tremor measurements across 12 sessions (4 weeks)—tremor scores showed a median reduction of more than 20%. A GLMM analysis indicated a significant dependence of tremor score on session progression (p = 0.002). The bradykinesia scores did not change significantly.

**Figure 2:**
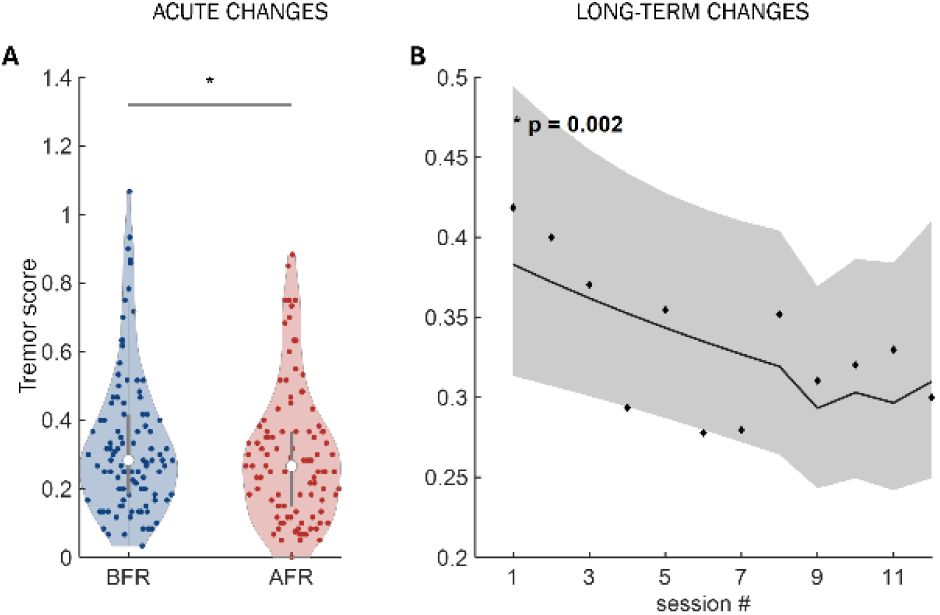
Shows changes in the tremor scores. (A) Acute change, (B) GLMM fit for changes over 12 sessions.

### Immediate changes in the LFP spectral features

Table 1 shows the changes in the spectral features such as total power, alpha, low-beta, high-beta frequency bands, and features derived from the FOOOF algorithm – ap_exp, and peak power. On the dorsal side of STN, we saw a median ∼8% increase in the low-beta band of the spectrum. We did not observe any significant changes in other features of the spectrum. On the ventral side, we observed a minimal increase (< 1%) in the low-beta band power. The ventral side also showed an increase (∼3%) in the median aperiodic exponent (ap_exp).

**Table 1:** The statistical description of individual features changes in immediate response to the biking exercise intervention.

| Feature | median (BFR) | IQR | Median (AFR) | IQR | Pval (WSRT, bonferroni corrected) |
| --- | --- | --- | --- | --- | --- |
| Dorsal (N = 21) |  |  |  |  |  |
| Total power | 23.99 | 10.65 – 37.34 | 24.52 | 11.77 – 37.26 | 0.44 |
| Alpha | 2.69 | 1.24-4.13 | 2.78 | 1.34-4.20 | 0.46 |
| Low - beta | 3.8 | 0.91-6.68 | 4.1 | 1.2-7 | **0.012** |
| High - beta | 6.53 | 3.15-9.92 | 6.72 | 3.2-10.25 | 0.26 |
| Aperiodic exp | 1.35 | 1.14-1.56 | 1.35 | 1.14-1.58 | 0.42 |
| Peak power | 0.57 | 0.44-0.69 | 0.55 | 0.43-0.67 | 0.09 |
| Ventral (N = 8) |  |  |  |  |  |
| Total power | 20.98 | 16.41 - 25.55 | 21.9 | 17.51-26.28 | 0.78 |
| Alpha | 2.6 | 2.06-3.12 | 2.65 | 2.04-3.26 | 0.51 |
| Low - beta | 3.24 | 2.04-4.43 | 3.25 | 1.59-4.91 | **0.015** |
| High - beta | 4.8 | 0.88-8.72 | 4.9 | 1.54-8.25 | 0.17 |
| Aperiodic exp | 1.21 | 1.07-1.35 | 1.25 | 1.11-1.39 | **0.018** |
| Peak power | 0.49 | 0.37-60 | 0.47 | 0.35-0.59 | 0.13 |

**Table 2:** SStr Estimate 1 used to assess causality between the dorsal and ventral systems.

| Condition on Strength<br>$\frac{AS_{ij}}{AS}$ | Condition on Cohesion | Inference |
| --- | --- | --- |
| Significant | $CS_i < CS_j$ | $S_i \rightarrow S_j$ |
| Significant | $CS_i \approx CS_j$ | $S_i \leftrightarrow S_j$ |
| Not Significant | $CS_i \approx CS_j$ | $S_i \vee S_j$ |
| Not Significant | $CS_i \neq CS_j$ | $S_i \perp S_j$ |

### Long-term changes in the LFP spectral features

*Long-term power changes* – Figure 3(A-D) present the results of a GLMM analysis of the data collected across 12 exercise (biking) sessions from electrodes placed in the dorsal and ventral regions. The X-axis represents session numbers, indicating the progression of the sessions, while the Y-axis shows corresponding power values for each of the electrode placements. The scatter plot shows the mean values of the spectral features of electrodes for each session. The dark solid line depicts the fitted mixed-effects model, capturing overall trends while accounting for fixed and random effects, with the light shaded region represents the confidence interval. Figure 3(A) shows that the total power in the dorsal region increases systematically with progressing session count (p = 0.002). On a finer scale, alpha power shows an increasing trend on the dorsal side (*p* = 0.004), but no trend on the ventral side. Low-beta (Figure 3C) and high-beta power (Figure 3D) both exhibit an increasing trend in the dorsal region (*p* = 0.003 and 0.004, respectively), and the ventral side displays no such trend.

**Figure 3:**
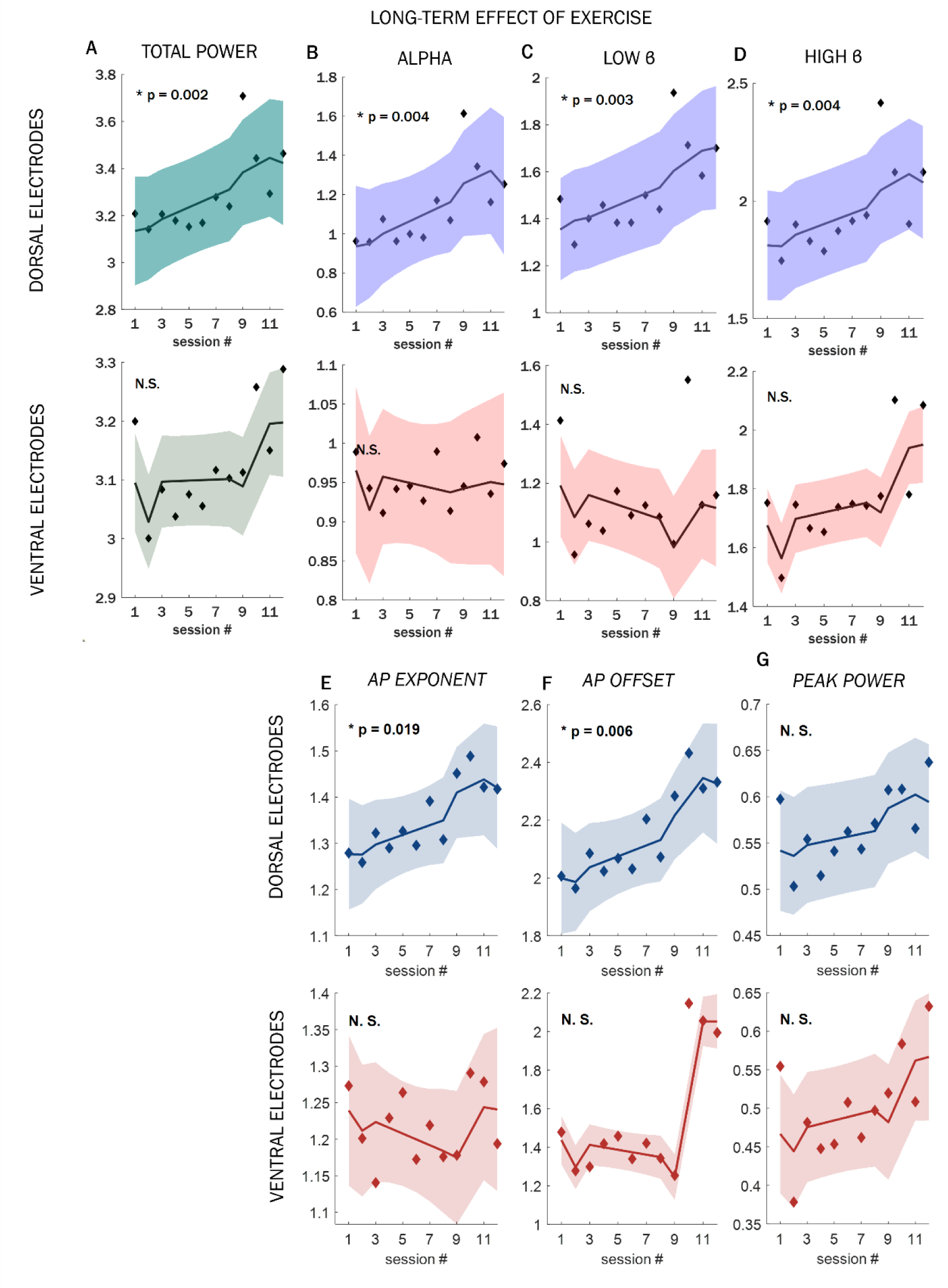
Plot of GLMM with session number as the fixed effect and the response variable shown alongside the GLMM fit for bot h ventral and dorsal regions for (A) total power, (B) alpha band, (C) low-beta band, (D) high-beta band, (E) aperiodic exponent, (F) aperiodic offset, and (G) peak power.

*Changes in the FOOOF features* – Similar to changes observed in canonical frequency band power, the background signal—characterized by the aperiodic exponent (ap_exp) and aperiodic offset (ap_off)—showed an increasing trend as the sessions progressed (Figure 3E-F). A similar pattern was also observed for peak power values (Figure 3G). Although the changes did not reach statistical significance, they exhibited a consistent upward trend over the course of the exercise sessions. This aligns with our previous findings, which also demonstrated increases in the aperiodic exponent and peak power over time. However, with the implementation of more advanced methods like GLMM to analyze trends in FOOOF features, we find that while the peak power follows a similar trend, its change is not statistically significant (Joshi et al., 2024).

### Changes in intra-STN communication and causality estimation

*Long-term changes in imaginary part of coh*erency – Figure 4A illustrates the long-term changes in iCOH, presenting the average pre-exercise (cycling) iCOH from the first two sessions compared to the last two sessions. While no significant long-term change is observed in the low-beta iCOH (WSRT, p = 0.38), the high-beta iCOH exhibits a significant increase over time (WSRT, p < 0.001). Figure 4B presents a representation of the session-wise increase in iCOH values between the dorsal and ventral regions of the STN.

**Figure 4:**
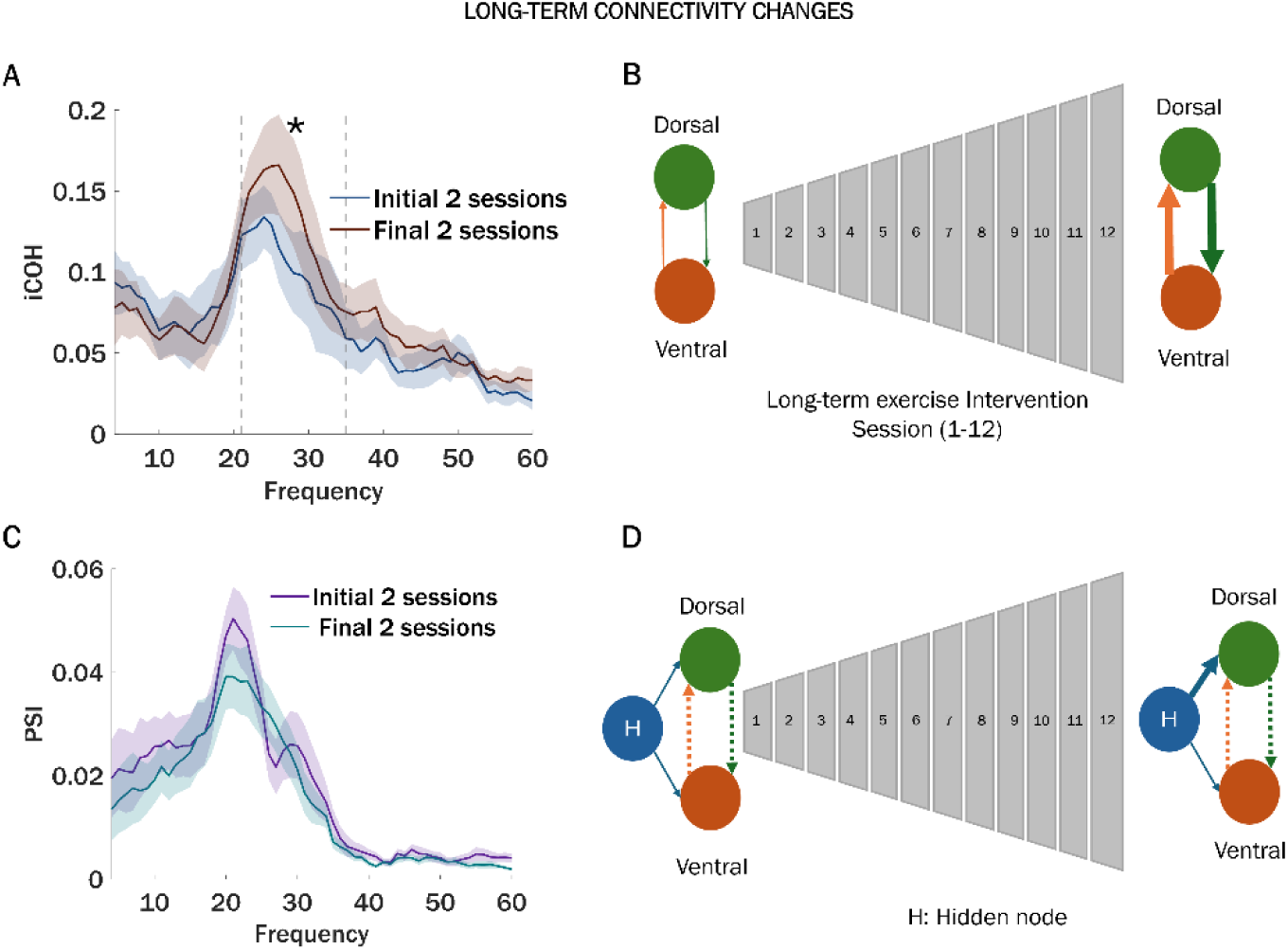
(A) iCOH ± SEM values for the 4-60 Hz frequency band across eight dorsal-ventral electrode pairs during the first two and last two sessions, before and after biking. (B) presents a schematic representation of the session-wise increase in iCOH values between the dorsal and ventral regions of the STN. (C) Displays the difference in pre-biking PSI values between the initial two sessions and the final two sessions of the intervention. (D) displays the findings of the SStr, showing a hidden node driving the dorsal and ventral STN in response to biking.

*Long-term changes in PSI* – Long-term changes in PSI are shown in Figure 4C. Using the jackknife method, PSI between 4 and 35 Hz was found to be statistically significant; however, the values remained below 0.1, indicating very low overall PSI. The PSI over this frequency range demonstrates no change in alpha, low-beta or high-beta frequency ranges following the long-term exercise intervention when comparing the values between the initial and final 2 exercise sessions.

*Analysis of the causal relationship between dorsal and ventral regions* – Evaluating Estimate 1 for Causality includes the assessment of whether 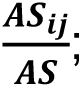 i.e.; the inter-system connectivity/total system connectivity is significant, and then a comparison of the subsystem cohesions: (i) If 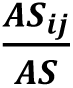 is significant, the subsystem cohesions (self-connectivity) are compared and the direction of causality is inferred accordingly to Figure 5A; else, (ii) If 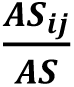 is not significant, then (a) If the subsystem cohesions are nearly equal, this indicates that the subsystems are driven by a hidden driver, or (b) If the subsystem cohesions are not equal, this indicates that the subsystems are independent.

According to (Sinha and Loparo, 2021), significance of 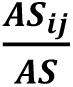 is determined either through (i) a comparison with the distribution of measures repeatedly generated under known experimental conditions or based on surrogate data analysis. Both of these methods for determining significance require a sufficient amount of experimental data. In this study, significance is assessed simply based on the fact that 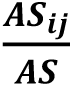 has a maximum value of 1 and a minimum of 0, therefore values < 6% (0.06) are considered not significant for this analysis.

The time-series of LFP recorded data from the dorsal and ventral regions before and after exercise were analysed using SStr where the nodes correspond to the positive and negative values of the dorsal and ventral LFP data connected to nodes that represent the temporal sampling points of the LFP data. The results in Table 3 use Estimate 1 (see Table 2) computed for all experiments across the 12 sessions, independently for data recorded before and after each exercise session. The analysis indicates that 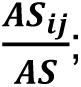 i.e., the inter-system connectivity/total system connectivity is not significant across all studies and exercise sessions with a maximum value of 6% (0.06). The overall trend in the relative values of dorsal and ventral subsystem cohesions that are within the range 0-2 before and after exercise indicates the existence of a hidden driver that is simultaneously influencing both the dorsal and ventral LFP time-series data. It is noted that the cohesion measure for a subsystem (S1) is the aggregate of arc system measures across self-connecting arcs within S1. Arc system measures have range of 0-1. For this analysis, cohesion measure has a range of 0-1, so with two subsystems defining the dorsal-ventral system, the range of the cohesion measures for the dorsal-ventral system is 0-2.

**Table 3:**
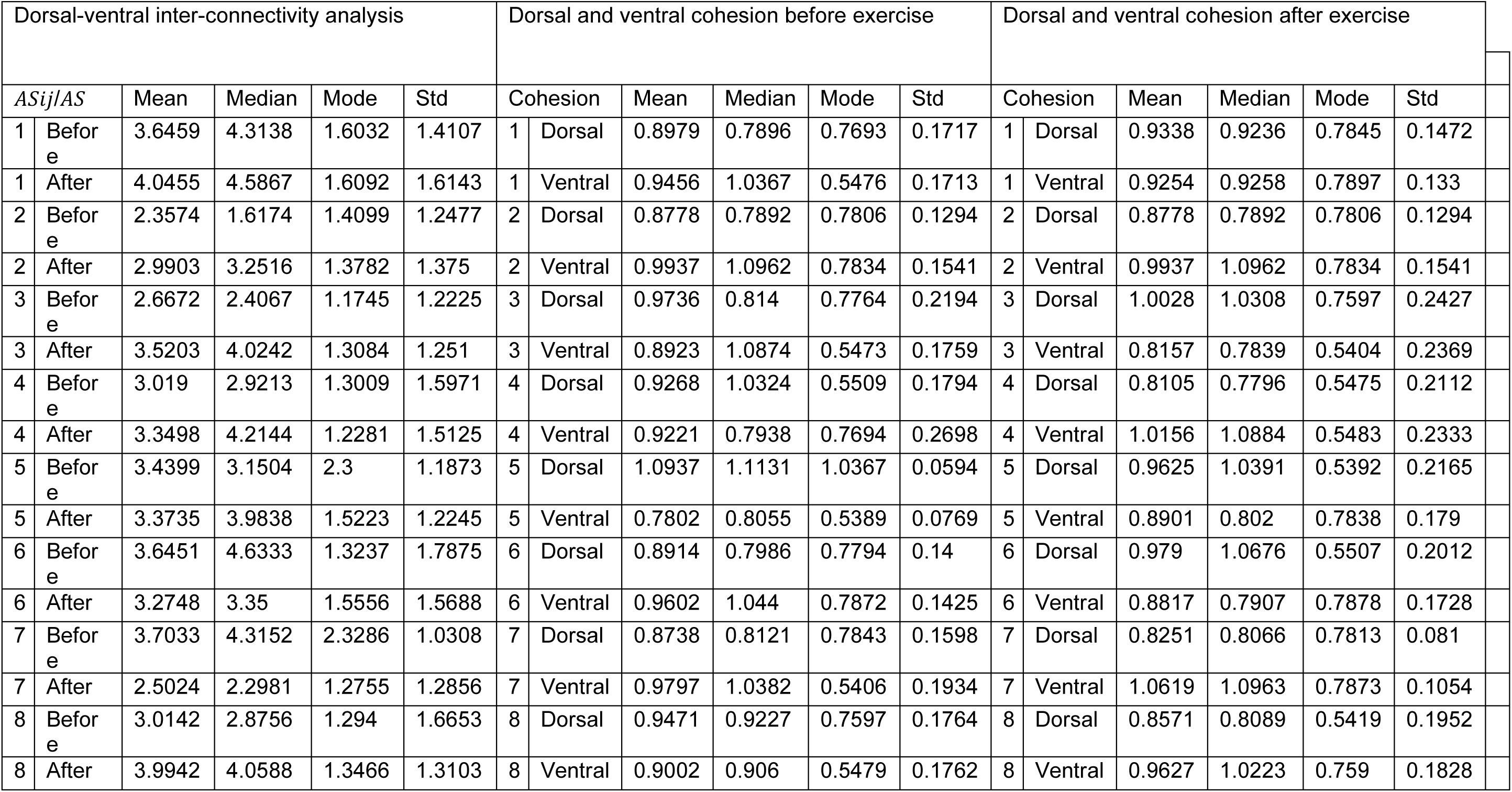
The results of SSTr method.

In conclusion, the general trend obtained from Estimate 1 as observed across exercise sessions and across subjects supports the hypothesis that inter-system (dorsal - ventral) interaction is driven by a hidden driver. Figure 4D presents a schematic summary of this finding. The purple hidden node (H) drives both the dorsal and ventral regions of the STN. Consequently, although there is no direct connection between the dorsal and ventral subregions, their activity appears correlated due to shared input from the hidden node. From available data, however, it is not possible to comment on feature or location of the hidden node.

## Discussion

The study aim is to investigate the short-term and long-term electrophysiological changes in the STN of patients with PD in response to a 12-session cycling exercise intervention. Our findings indicate that electrodes positioned in the dorsolateral region of the STN show the most prominent response to exercise. Over the length of the intervention, there was an increase in the STN activity, not only limited to the widely studied beta band, but also in the alpha band.

### Motor symptoms

Although the initial tremor score was low (<1 on a 0–4 scale) due to symptom assessment with DBS remaining active, exercise led to further improvement, with long-term changes being more pronounced than acute effects. In contrast, bradykinetic scores did not exhibit significant changes, suggesting that dynamic biking may specifically target tremor-related neural circuitry or that bradykinetic symptoms may require a longer or alternative intervention. Consistent with these findings, previous studies on PD patients without DBS have demonstrated improvements in upper extremity motor symptoms following the same 4-week intervention. The observed improvements in upper extremity symptoms in response to a lower extremity exercise regimen suggest the involvement of central nervous system mechanisms, with particular interest in changes within the STN, which are the focus of the present study.

### Changes in Spectral features of LFP

In the acute timeframe, low beta power increased significantly after an exercise session. This suggests enhanced communication within the low-beta frequency range compared to other frequency bands. The low-beta band is known to respond to dopamine medication (Mathiopoulou et al., 2024). However, most participants are on extended-release carbidopa/levodopa, which typically maintains stable drug levels for approximately 6 hours following intake at time t = 0 (Margolesky and Singer, 2018). All our experiments were conducted in the middle of this time window, further keeping the timing of biking and dopamine intake consistent across trials. With this information, we are positive that the concentration of dopamine remained stable when the pre-biking and post-biking data was collected. Given this, the increase in low-beta power likely reflects mild, exercise-induced modulation rather than fluctuations in medication. In contrast, no significant changes were observed in the alpha or high-beta bands, suggesting that the acute effects of exercise may be frequency-specific, with low-beta showing the most sensitivity.

Over the long term, all sub-bands along with the total power showed significant increases in power with progressive sessions, suggesting neuroplastic adaptations. In contrast, on the ventral side remains unchanged in response to exercise. Spectral parameters derived from the FOOOF algorithm suggest that these changes, particularly in the beta bands, result from the strengthening of neural oscillators and alterations in background signals. In PD literature, most interventions are designed to reduce hypersynchrony in the STN, which is frequently associated with symptom improvement (Brown, 2007; Jenkinson and Brown, 2011; Kuhn et al., 2008; Kühn et al., 2006; Mathiopoulou et al., 2024). Studies have documented reductions in STN beta synchrony following interventions such as levodopa therapy, evidenced by decreases in absolute (Sumarac et al., 2024), relative (Kühn et al., 2006; Neumann et al., 2017), or oscillatory beta-band power (Liu et al., 2024; Mathiopoulou et al., 2024). While not all studies have accounted for the precise location of the recording electrodes, this reduction in beta activity is widely regarded as a key indicator of therapeutic efficacy. Moreover, reductions in beta-band power are often correlated with improvements in a subset of PD symptoms. However, our findings reveal an unexpected increase in beta power following exercise, with a well-established intervention for alleviating PD symptoms (Alberts et al., 2016; Kim et al., 2024; Ridgel et al., 2015; Ridgel and Ault, 2019). This observation is particularly intriguing as it contradicts the prevailing expectation of beta power reduction. These results suggest the possibility of mechanisms beyond the basal ganglia-cortical pathways.

The changes in the motor outcome are more prominent in the tremor subtype compared to the bradykinesia subtype. This may indicate that the neural substrate involved in the generation of tremor could be a potential driving factor of the changes in the STN. Based on the recent literature, one such potential structure implicated in this process could be the cerebellum (Hallett, 2014; Li et al., 2023; Wu and Hallett, 2013). Evidence from mouse models and human fMRI studies indicates that the cerebellum is responsive to exercise interventions, such as treadmill walking (Ding et al., 2022; Li et al., 2022). The cerebellum, which is intricately connected to the motor cortex and plays a critical role in movement and coordination, has recently been identified as having bidirectional, bi-synaptic connectivity with the basal ganglia. This connectivity positions the cerebellum as a compelling candidate for further investigation into its role in exercise-induced changes in neural activity and PD symptomatology.

In the classical basal ganglia model, the STN primarily receives input from two connected key nuclei. It receives inhibitory input from the globus pallidus externus (GPe) via GABAergic projections as part of the indirect pathway (Smith et al., 1990) and excitatory input from the motor cortex through the hyperdirect pathway (Nambu et al., 2002). The dorsal region of the STN is specifically connected to the premotor cortex, which is also known to drive high-beta activity via the hyperdirect pathway. Hence, one of the possible explanations for the increase in STN power could be related to the strengthening of the hyperdirect pathway.

Another intervention which is known to increase the STN is the anticholinergic treatment of PD that is known to help with tremors and rigidity (Priori et al., 2004). This provides an example of the case where the beta power increases, but the symptoms reduce. While there isn’t enough evidence to establish a connection between exercise and anticholinergic behavior, this fact may suggest investigating different types of neurotransmitters available and other regions that could be influencing the basal ganglia activity indirectly in the circuitry to identify the reason behind an increase in the beta band power.

The dynamic biking exercise intervention engages the rider in a continuous feedback loop, where internal model predictions are updated based on sensory input to adapt to the bike’s motor response (Francis and Wonham, 1976; Kawato, 1999; Welniarz et al., 2021). In this process, proprioception is one of the senses that participates in feedback. Peripherally, proprioceptive signals interface with the brain through second-order neurons via two primary pathways: the dorsal column–medial lemniscus pathway (Gilman, 2002; Wang et al., 2021), which relays information to the thalamus, and the dorsal and ventral spinocerebellar tracts (Matsushita and Hosoya, 1979; Xu and Grant, 2005), which transmit input to the cerebellum. Both the thalamus and cerebellum interact directly or indirectly with the basal ganglia circuitry possibly via caudate nucleus and substantia nigra (Nagy et al., 2006). Therefore, repeated engagement in cycling may facilitate communication between the basal ganglia–thalamo–cortical and cerebello–thalamo–cortical circuits through proprioceptive feedback, potentially promoting neuroplastic changes. However, the precise nature of these interactions remains unclear and warrants further specific investigation into the effects of proprioceptive changes on STN-LFPs.

### Changes in coherency and PSI within the STN

The imaginary part of coherence revealed distinct immediate effects during initial and later sessions. Over the long term, exercise resulted in an overall increase in high-beta coherence within the STN. To determine whether these coherence changes originated from within the STN or were influenced by external sources, the PSI that measures the directionality of information flow between the dorsal and ventral regions was calculated. PSI values remained below 0.1, indicating minimal crosstalk between these regions, consistent with the limited presence of interneurons within the STN. This aligns with findings by (Steiner et al., 2019) suggesting that STN neurons function as independent processing units, with their activity primarily driven by upstream structures. It suggests that dorsal and ventral electrodes are likely to record distinct oscillatory activity driven by external sources. Given STN’s crucial role in motor circuitry, the minimal crosstalk implies that changes in the imaginary part of coherence are more likely mediated by external structures rather than intrinsic STN communication. Because PSI values indicate negligible internal communication changes within the STN, these findings suggest that an external structure is modulating interactions with both dorsal and ventral regions. Our preliminary study of the causal relationship between the dorsal and ventral regions also points to an external source (hidden driver) that is simultaneously impacting the temporal patterns of the LFP time-series in the two regions. Furthermore, the observed neuroplastic changes appear to influence the dynamics of this external communication.

Our preliminary study investigating causal relationships between the LFP time-series recorded from the dorsal and ventral regions indicated non-significant inter-connectivity between the two regions, and nearly equal dorsal and ventral cohesion measures indicative of a hidden driver that influences both regions. It was hypothesized that the exercise intervention was potentially a hidden driver, but our preliminary results indicate similar inter-connectivity and cohesion measures before and after exercise, with no distinctive trends over the course of the exercise sessions. The cerebellum is a candidate for the hidden driver, and additional studies are required to investigate this further. Due to the complexity of the basal ganglia and movement control circuitry, a comprehensive understanding of these dynamics will require accounting for changes across multiple interconnected regions. Without such considerations, this remains a multivariable problem with a single equation.

## Conclusion

We investigated the effects of dynamic cycling intervention in individuals with PD by analyzing LFP signals recorded from the STN before and after exercise, across 12 sessions conducted over a 1-month period. The goal was to explore the neural mechanisms underlying observed improvements in PD symptoms, particularly tremors. While only minimal changes were observed in the low-beta band acutely following exercise, we found a robust long-term increase in overall STN power with continued intervention. Additionally, power within specific frequency bands such as alpha, low-and high beta also increased, suggesting exercise-induced neuroplasticity within the STN. Connectivity analysis revealed no direct interaction between the dorsal and ventral subregions of the STN; however, both regions displayed coherent activity, implying the influence of a common external driver. Using the SSTr method, we were able to identify and demonstrate the presence of this hidden driver.

## Data Availability

All data produced in the present study are available upon reasonable request to the authors

## Acknowledgements

Our gratitude extends to the Medtronic, Minneapolis, MN, USA team particularly James Adler and Robyn Whipple, and Adam Matthews for their invaluable support in steering us through the data collection and raw data interpretation process. We also extend our thanks to the whole team at InMotion, Beachwood, Ohio, USA for providing us with the support and facility to conduct our experiments. The study was supported by United States Department of Veterans Affairs grant I01RX003676 (Shaikh). Shaikh was supported by grants from American Parkinson’s Disease Association (George C. Cotzia Fellowship); and has received Penni and Stephen Weinberg Chair in Brain Health.

## Author’s Roles

P.J.: Study design, data collection, data analysis, writing-original draft. L.M.F: data collection, writing-review and editing.

B.E.S: data collection, writing-review and editing.

C.W.K: participant recruitment, data collection, writing-review and editing. A.G: Data analysis, editing.

K.A.L Causality analysis, writing-review and editing.

A.K.S. Causality analysis

A.L.R: project administrations and supervision, writing-review and editing.

A.G.S: Study design, data collection, participants recruitment, project administrations and supervision, Writing-original draft, review and editing

